# Privacy-Preserving Distilled Large Language Models Enhance Multimorbidity Scoring

**DOI:** 10.64898/2026.09.19.26363476

**Authors:** Raghav Awasthi, Yihe Yang, Mengxuan Li, Xiaofeng Zhu

## Abstract

The truthful use of large language models (LLMs) is a growing challenge in safeguarding sensitive patient data from leakage. We introduce and evaluate a privacy-preserving knowledge distillation framework for LLM-based clinical modeling, using multimorbidity scoring as a healthcare task. Although LLMs can encode rich clinical knowledge and improve upon traditional rule-based comorbidity scoring, their direct evaluation on large-scale biobank data remains constrained by patient privacy. In our framework, multimorbidity reasoning is distilled from state-of-the-art LLM teacher models into compact student models (CoLLMs) using synthetic cohorts that preserve UK Biobank distributions, without exposing real patient data. This approach achieves high-fidelity knowledge transfer (Spearman ρ = 0.75–0.89). Independent LLM-as-Judge evaluation confirms the clinical significance of the distilled knowledge and reveals substantial variability among teacher models. When applied to real UK Biobank data, CoLLM-derived multimorbidity scores improve survival prediction (C-index up to 0.91) and exhibit higher SNP heritability (h^2^ ≈ 0.05). Our work establishes a trustworthy, privacy-compliant pathway for large-scale healthcare applications of LLMs.

## Main

More than one-third of adults worldwide live with multimorbidity^1,2^. Multimorbidity, defined as the presence of multiple diseases within an individual substantially affects disease severity and overall health outcomes^3–5^. The terms comorbidity and multimorbidity are sometimes used interchangeably; however, they differ in their clinical focus. Comorbidity refers to the coexistence of additional diseases centered around a primary disease, whereas multimorbidity refers to the presence of multiple diseases without any single primary condition. Both comorbidity and multimorbidity correlate with increased all-cause mortality, higher hospitalization rates, longer hospital stays, higher healthcare utilization, and reduced quality of life^2,6,7^. The COVID-19 pandemic highlighted the impact of multimorbidity burden, as individuals with comorbidities faced increased risks of severe infection, intensive care admission, and mortality^8–10^. Prior research has shown that multimorbidity plays a central role in major causes of death worldwide, including cardiovascular disease and cancer^11–14^. This makes the accurate measurement of multimorbidity essential for prioritizing interventions, such as preventive measures, vaccination, early treatment, and hospital resource allocation.

Several rule-based approaches, including the Charlson comorbidity index (CCI) and the Elixhauser comorbidity index (ECI) have been widely used to quantify multimorbidity risk^15–17^. These scores systematically rank disease severity based on the assumption that more severe diseases contribute higher weights, whereas less severe diseases contribute lower weights. Although these indices follow established clinical conventions and are simple to apply, they face important limitations. One major limitation is the limited disease coverage; both the ECI and CCI only cover less than 1% of all defined ICD-10 codes, leaving out many clinically relevant diagnoses and interactions^18^. Another limitation is that they do not capture the non-linear interplay among diseases. In addition, they fail to account for diverse socio-economic status and other electronic health record (EHR) information that may influence comorbidity risk^18^.

LLMs have shown potential to solve complex healthcare tasks by addressing complex clinical tasks that require contextual reasoning^19–22^. LLMs are trained on large datasets, and their complex architecture, which is aligned with feedback-driven optimization strategies, enables them to capture rich semantic and contextual representations of language. Recent literature also confirms that LLMs have achieved near-expert performance across a range of clinical applications, including answering USMLE-style examination questions with over 90% accuracy, summarizing lengthy clinical notes, supporting diagnostic reasoning, preliminary assessment of patients, and providing healthcare guideline-based decision support^21,23–25^. This performance highlights the promise of LLMs for a new decision support paradigm in health informatics.^20^ However, building widespread trust in LLMs requires rigorous and transparent evaluation. A basic form of evaluation includes directly providing patient information to an LLM and evaluating whether it produces clinically appropriate outputs. A major and often under explored challenge in evaluating LLMs is data privacy: most state-of-the-art LLMs such as Gemini, Deepseek, GPT are closed-source and accessible only through external APIs, making them unsuitable for direct use with sensitive datasets governed by strict data-use agreements^26–28^. For example, UK Biobank policy does not allow researchers to incorporate participant level data into publicly available LLMs^29^. Furthermore, there remains active debate in the scientific community regarding whether LLMs demonstrate true reasoning in biomedical contexts, as evidence suggests that their outputs often reflect pattern recognition rather than true reasoning^30–33^.

In this study, we followed an evaluation strategy motivated by a knowledge distillation framework to address privacy constraints. Knowledge distillation is a machine learning technique in which the capabilities of a large pre-trained “teacher model’’ are transferred to a smaller, more efficient “student model’’^34^. Prior literature suggests that student models can sometimes even outperform their teacher models^35,36^. This framework allowed us to avoid exposing real patient data to closed-source third-party LLMs. Instead, we generated synthetic patient profiles based on UK Biobank distributions, used these to evaluate teacher LLMs, and trained student models ***CoLLMs (Comorbidity Scoring via Large Language Models)*** to mimic teacher LLM derived scores for multimorbidity scoring. The high-fidelity CoLLM student models were applied to real UK Biobank data to generate multimorbidity scores, which were evaluated through survival analysis, genome-wide association studies, and associations with polygenic risk scores (PRS). We additionally used an LLM-as-Judge framework to qualitatively evaluate both teacher LLMs and CoLLMs, assessing their clinical alignment, reliability, and safety. Our study makes four key contributions: (1) We developed and validated CoLLMs using 50,000 synthetic patient profiles derived from more than 400,000 UK Biobank records, encompassing diverse diseases and demographics. (2) We demonstrated that CoLLM-derived scores outperform existing comorbidity indices (CCI and ECI) in survival analysis and reveal meaningful genetic associations. (3) We showed that our distilled models retain approximately 90% concordance with teacher LLMs while achieving faster inference and full privacy compliance. (4) We conducted qualitative evaluation of both teacher LLMs and student CoLLMs using an independent LLM-as-Judge approach to ensure meaningful clinical behavior and performance.

To support clinical translation and usability we also provide an open-source web application for real-time CoLLM-based risk prediction (https://zhulab-collm.streamlit.app/), along with all model weights and training code. This framework offers a generalizable approach for applying advanced AI to large-scale health research while preserving patient privacy and scientific rigor.

## Results

### Workflow for the Development and Evaluation of CoLLM

CoLLM is a LLM distilled neural network designed to generate multimorbidity risk scores from an individual’s ICD-10 code lists and demographic variables (age and gender) (**see Fig. 1)**. The model was explicitly developed to mimic the behavior of LLMs while mitigating privacy concerns associated with exposing sensitive health data to third party APIs.

**Figure 1:**
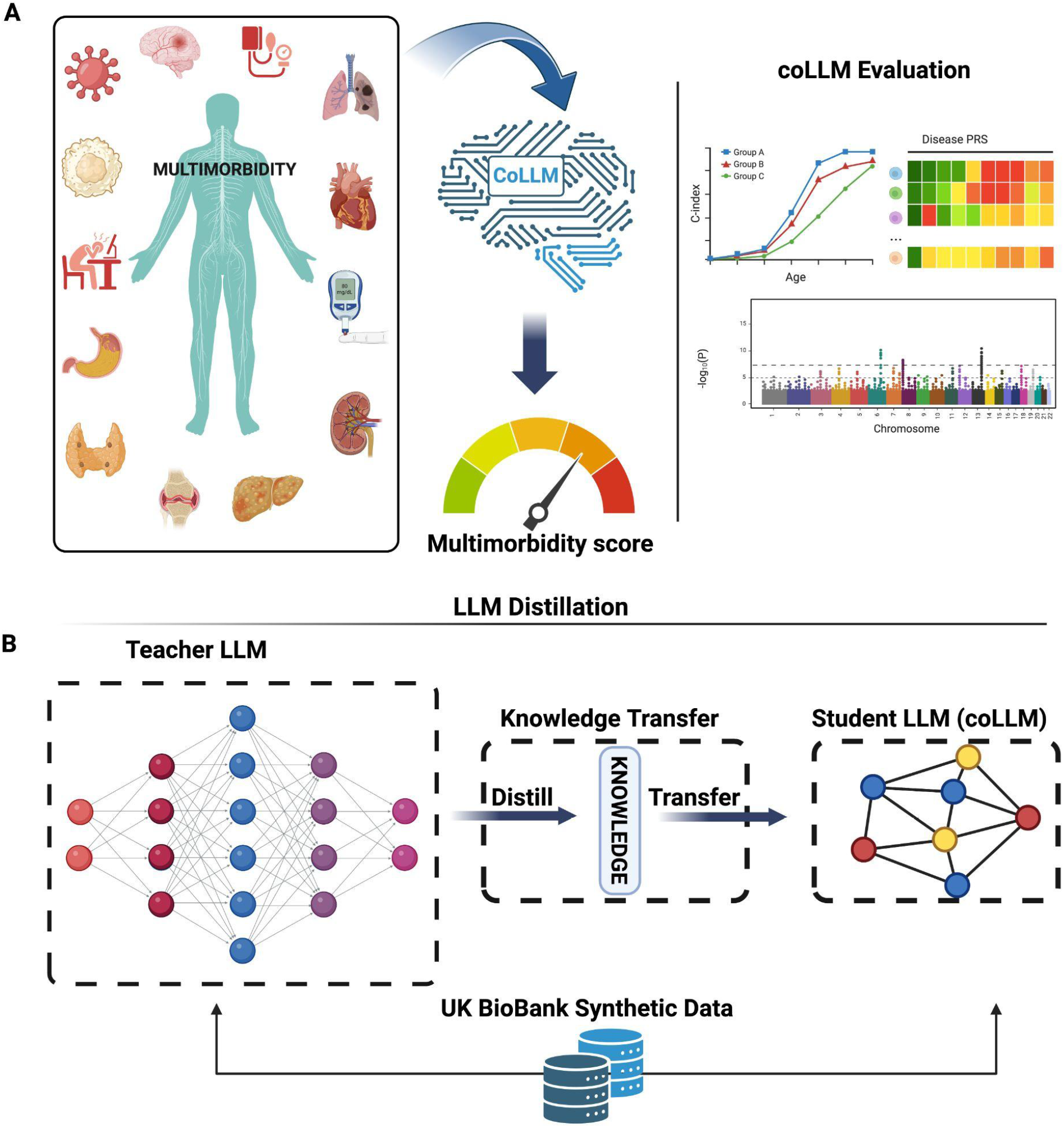
Overview of the CoLLM framework for multimorbidity risk prediction and evaluation. (A) Flowmap of multimorbidity score calculation. The CoLLM model incorporates ICD-10 disease codes, age, and gender to generate multimorbidity scores. These scores are evaluated across diverse biological and clinical dimensions. Evaluation includes survival prediction, polygenic risk score correlations, and genome-wide association studies. (B) Conceptual diagram of the LLM distillation process. A large teacher LLM processes synthetic UK Biobank-derived data to generate multimorbidity scores. These scores are then used to train a smaller student model (CoLLM) using a knowledge distillation pipeline. This enables CoLLM to mimic the reasoning of large LLMs while maintaining computational efficiency and compliance with privacy regulations for large-scale health data.

To enable this, ICD-10 codes were grouped into 263 parent disease categories **Supplementary Data 1)** using UK Biobank (UKB) data from 439,221 participants. Synthetic datasets were generated using Conditional Tabular Generative Adversarial Networks (CTGAN)^37^ with 50,000 samples for training and 10,000 for testing. For each synthetic individual we used three state-of-the-art LLMs (GPT-4^38^, Gemini^39^, and DeepSeek^40^) (**detailed model specifications in Supplementary Data 6**) in a zero-shot prompting setup providing ICD-10 codes age and sex to derive comorbidity scores (**detailed prompt in Supplementary Note 2**).

These LLM-derived scores were used as comorbidity score labels as the supervised training data for CoLLM. The network was trained to predict comorbidity scores in four configurations: each LLM individually and the mean of all three models. Training was performed on 50,000 synthetic samples, with evaluation on 10,000 held-out samples using Pearson correlation.

Next, we conducted a qualitative evaluation of both teacher LLMs and student CoLLMs using an independent LLM-as-Judge framework. This assessment quantified the clinical Alignment, Score reliability, and Outlier and safety risk of all models on Likert-scale criteria (**details in Supplementary Note 3)**. Finally, CoLLM was applied to the full UKB dataset (N = 439,221) to generate distilled LLM comorbidity scores. These scores were then evaluated across multiple dimensions: survival prediction, genome-wide association studies (GWAS), and associations with disease-specific polygenic risk scores (PRS). CoLLM-derived scores were benchmarked against widely used comorbidity scores such as CCI and ECI.

### Comprehensive Comparison of Synthetic and Real Cohort Characteristics

The quality of synthetic data generated by the CTGAN was evaluated by benchmarking demographic and clinical characteristics against the original UK Biobank cohort (**see Fig. 2)**. To quantitatively assess the fidelity of the CTGAN-generated synthetic cohort, we used the Synthetic Data Vault (SDV) evaluation framework ^41^. SDV computes synthetic data fidelity scores by comparing how well the synthetic data reproduces univariate distributions, called the Column Shapes Score, and bivariate relationships between feature pairs, called the Column Pair Trends Score. It then aggregates these into an overall fidelity score. We found that the Overall fidelity was high, as reflected in the SDV evaluation metrics: the Column Shapes Score was 96.69%, the Column Pair Score was 93.99%, resulting in an overall synthetic data quality score of 95.34%. These scores indicate strong preservation of both individual variable distributions and joint relationships across features. Demographic characteristics were closely reproduced. Age showed high distributional similarity between real and synthetic cohorts, with a KSComplement (1− KS distance) of 0.927. Sex and ethnicity were also well preserved, with TVComplement (1 − Total Variation distance) scores of 0.776 and 0.876, respectively (Fig. 2A–C). At the diagnostic level, prevalence across 263 parent ICD-10 categories correlated strongly between the real and synthetic cohorts (Pearson r = 0.73, p < 0.001) **(see Fig. 2D)**. Comorbidity structure was evaluated using clinically relevant disease pairs by comparing the observed prevalence of each pair between real and synthetic cohorts (**see Supplementary Data 2**). Correlation was moderate (Pearson r = 0.58, p = 1.50e-03), suggesting that higher-order co-occurrence patterns among key comorbidities were broadly retained, although not as closely as single-disease prevalence (see Fig. 2E). Some differences in disease prevalence were also identified. Hypertension (73.1% in synthetic data versus 34.4% in real data) and falls (21.6% versus 9.9%; accidental falls) were over-represented. In contrast, general symptoms and signs (4.3% vs. 21.7%), arthrosis (5.1% vs. 21.1%), ischaemic heart disease (4.4% vs. 13.0%), and diabetes mellitus (2.7% vs. 9.8%) were under-represented. These discrepancies reflect established challenges in deep generative models **(Fig. 2F)**. We also evaluated attribute-disclosure risk using SDMetrics’ *DisclosureProtectionEstimate* treating each ICD-10 parent disease as the sensitive attribute and using demographic information (age, sex, ethnicity) as identifiers. Among the top five prevalent disease groups, disclosure-protection scores ranged from 0.77 to 1.00, all exceeding the baseline value of 0.50 **(Supplementary Data 8)**.

**Figure 2.**
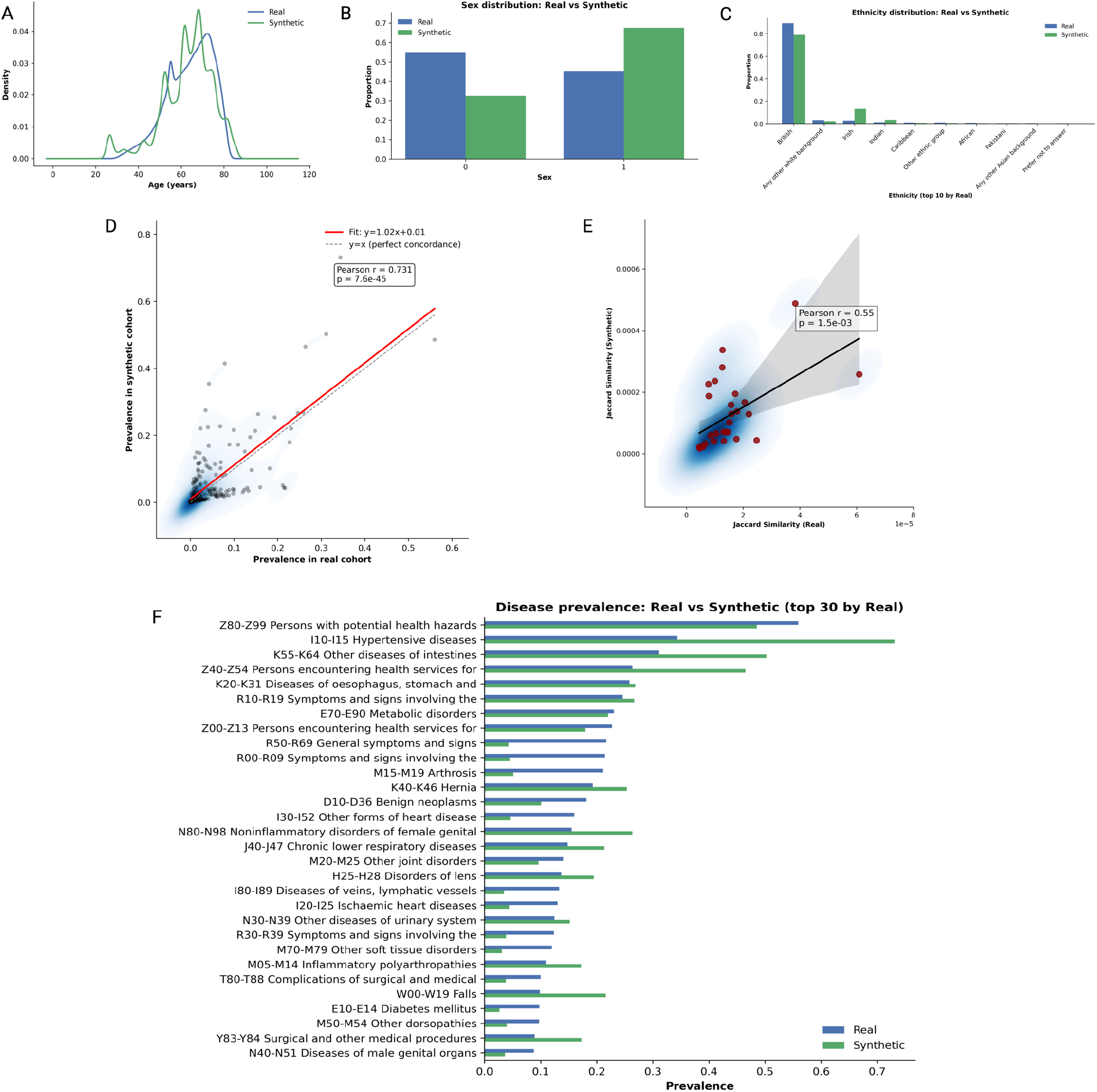
Evaluation of Synthetic Data Generation from UK Biobank using CTGAN. (A) Density plot of age distribution for both real and synthetic data. (B) Bar plot showing sex distribution (male and female) in real and synthetic data. (C) Bar plot showing ethnicity distribution in real and synthetic data. (D) Correlation of disease prevalence across all ICD-10 codes between real and synthetic data. (E) Correlation of pairwise disease co-occurrence represented by the Jaccard Index. (F) Top 30 prevalent diseases in the real data and their corresponding prevalence in both real and synthetic datasets.

To further assess prevalence-level fidelity, we quantified absolute prevalence errors across disease categories. Across ICD-10 parent disease categories, the mean absolute prevalence error was 0.0319, the median absolute error was 0.00794, and prevalence-stratified analysis showed that absolute error increased with disease frequency, from 0.0056 in rare conditions (<1%) to 0.1012 in common conditions (≥10%) **(Supplementary Figure 3)**.

### CoLLM achieves high correlation in mimicking LLMs

Four CoLLM networks were trained: three student models mimicking individual LLM outputs, and one ensemble model (LLM-Mean) mimicking the average output of all three LLMs. Each model was trained on 50,000 synthetic samples and evaluated on 10,000 held-out samples. All CoLLMs achieved strong correlation with their respective teacher models **(Fig. 3)**. Spearman correlation coefficients ranged from 0.75 for CoLLM-DeepSeek to 0.89 for CoLLM-Mean, with CoLLM-Gemini (0.80) and CoLLM-GPT-4o (0.77) **(Fig. 3A)**. Residual analysis showed minimal dependence between predicted values and residuals for all CoLLMs (|ρ| < 0.22), indicating that prediction errors were largely stable across the score range. CoLLM-Mean in particular exhibited near-zero correlation (ρ = 0.055), suggesting well-balanced errors with no systematic over- or underestimation across predicted multimorbidity levels **(Fig. 3B).**

**Figure 3.**
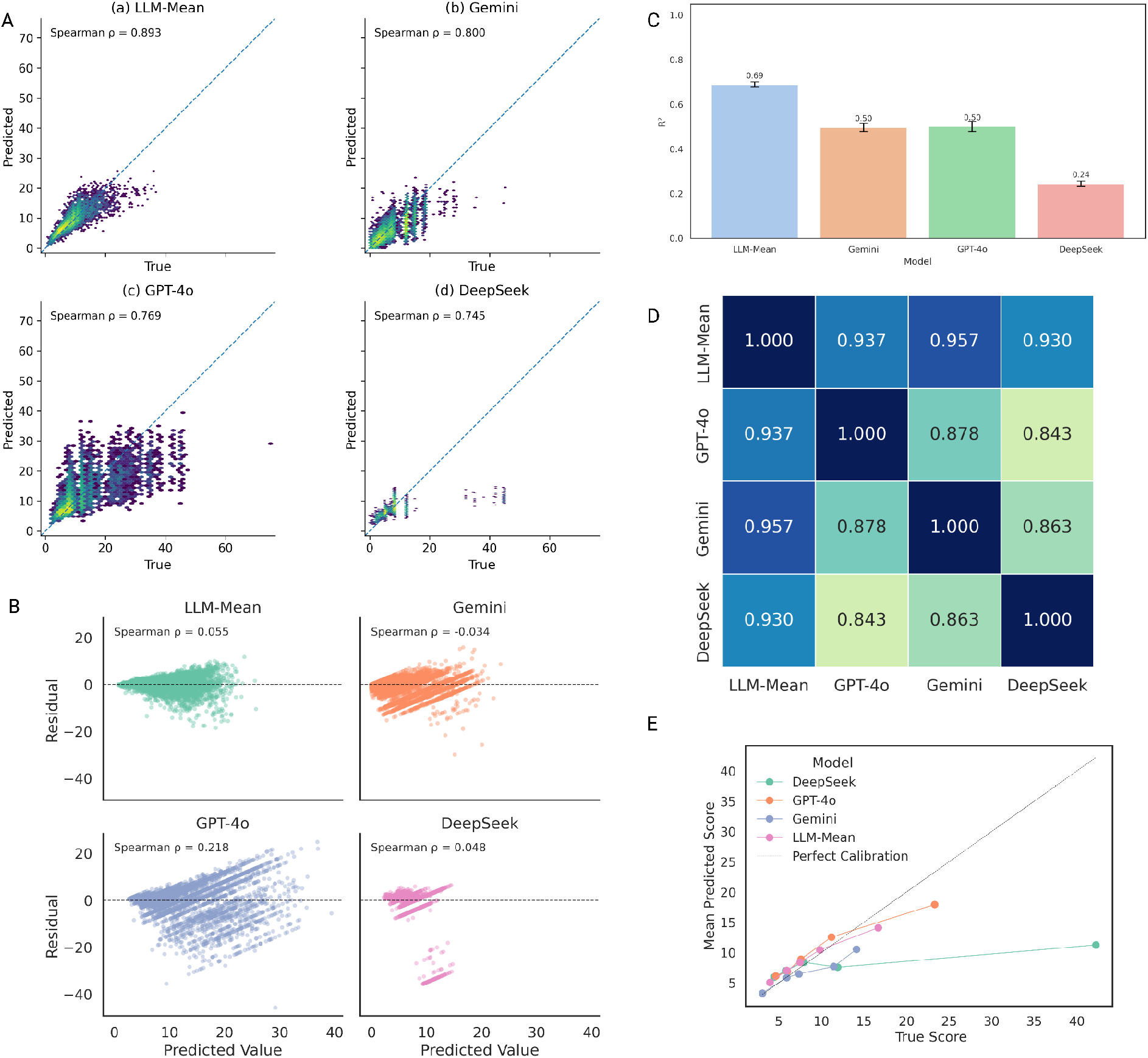
Evaluation of CoLLM’s ability to mimic multimorbidity score generation by Large Language Models. (A) Correlations between true LLM-derived multimorbidity scores and CoLLM-predicted scores on the test set. (B) Residuals of CoLLM-predicted scores. (C) Coefficient of determination (R^2^) values comparing LLM-derived and CoLLM-predicted scores. (D) Heatmap showing pairwise spearman correlations among CoLLM-predicted scores. (E) Calibration curve generated after dividing the true scores into 5 quantile bins and computing the mean CoLLM-predicted score within each bin. CoLLM-Mean most closely follows the identity line, indicating superior calibration relative to individual CoLLMs.

Model-specific R^2^ values further supported these results. LLM-Mean demonstrated the highest score (R^2^ = 0.69), outperforming individual CoLLMs (Gemini 0.50, GPT-4o 0.42, DeepSeek 0.24) **(Fig. 3C).** Pairwise spearman analysis indicated high cross-model consistency (r > 0.84), with the strongest correlation observed between CoLLM-Gemini and LLM-Mean (r = 0.96) **(Fig. 3D)**. The calibration curve shows that CoLLM-Mean most closely tracks the teacher LLM scores across all bins, staying nearest to the perfect-calibration line. In contrast, individual CoLLMs, especially DeepSeek, progressively underpredict higher multimorbidity scores, indicating increasing bias at the upper range **(Fig. 3E)**..

### CoLLM-derived multimorbidity scores demonstrate superior prognostic performance compared to traditional indices in the UK Biobank

We next applied CoLLMs to generate multimorbidity scores on real-world UK Biobank data and evaluated their prognostic utility against traditional indices for all-cause mortality risk discrimination (median follow-up 4.8 years [IQR 2.7–9.2]; n = 37,094 events) **(Fig. 4 A).** Across all age strata, CoLLM-based scores achieved higher and comparable C-indices than the CCI and ECI, with the LLM-Mean ensemble consistently demonstrating optimal performance.

**Figure 4.**
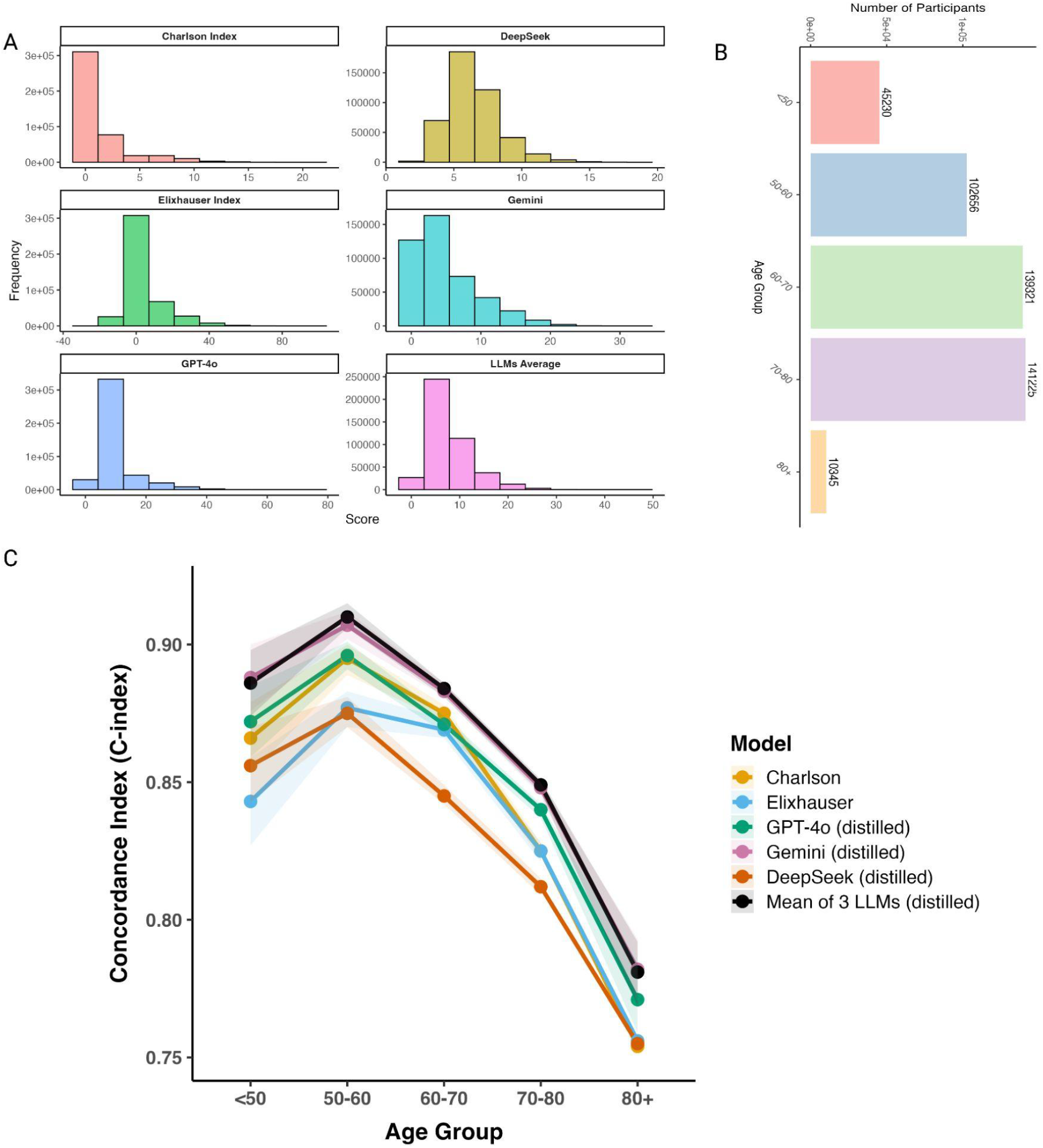
Survival analysis on UK Biobank data. (A) CoLLM-derived multimorbidity scores applied to UK Biobank participants. (B) Bar plot showing age group–wise distribution of individuals. (C) Concordance index (C-index) with 95% confidence intervals from Cox proportional hazards models using multimorbidity scores as predictors.

In participants aged 50–60 years **(Fig. 4B)**, LLM-Mean achieved a C-index of 0.910 (95% CI 0.906–0.915), significantly outperforming both CCI (0.895 [0.889–0.900]) and ECI (0.877 [0.870–0.883]). Comparable improvements were observed in the 60–70 year cohort, where LLM-Mean reached 0.884 (0.881–0.886) versus Charlson (0.875 [0.872–0.878]) and Elixhauser (0.869 [0.866–0.873]). Performance gains persisted even in the oldest age stratum (≥80 years), where discriminative ability declined across all models: LLM-Mean maintained superior performance (0.781 [0.769–0.792]) relative to Charlson (0.754 [0.741–0.766]) and Elixhauser (0.756 [0.743–0.769]) **(Fig. 4C)**.

Among individual CoLLMs Gemini consistently outperformed GPT-4o and DeepSeek (C-index for 50–60 years: 0.907 versus 0.896 versus 0.875, respectively), though it remained comparable to the LLM-Mean. These findings demonstrate that CoLLM-derived scores particularly the Gemini and LLM-Mean provide enhanced survival risk stratification across age groups compared with established comorbidity indices, highlighting their potential for population-scale prognostic applications. Also, The time-dependent AUC analysis showed consistent discriminatory performance across 1-, 3-, and 5-year follow-up periods, supporting the model’s ability to stratify mortality risk over time; these results are reported in **(Supplementary Data 10)**

### Genome-wide association analysis reveals that CoLLM-derived multimorbidity traits capture meaningful genetic architecture

We performed genome-wide association studies (GWAS) using the Genome-Wide Robust Analysis for Biobank Data (GRAB) method^42,43^ across all six multimorbidity traits: traditional indices (CCI and ECI) and four LLM-derived scores (LLM-Mean, GPT-4o, Gemini, and DeepSeek). GRAB was selected for its ability to model ordinal phenotypes. To enhance statistical power, each trait was stratified into four ordinal groups representing increasing multimorbidity severity, enabling the mixed-model framework to effectively capture polygenic effects across the disease burden spectrum **(Supplementary Table 4)**.

Genome-wide analyses of CoLLM-derived multimorbidity traits revealed a combination of shared and model-specific loci (**Supplementary Table 3, Fig. 5A**). The most consistent and significant (p < 5 × 10−8) association was observed at the *HLA-DQA1/DQB1* region on chromosome 6, a canonical immune locus showing strong signals across multiple models—CCI (−log_10_P = 35.93), LLM-Mean (−log_10_P = 10.54), Gemini (−log_10_P = 12.09), and GPT (−log_10_P = 8.07). The HLA region plays a central role in immune regulation and is well known for its association with autoimmune diseases such as coeliac disease, type 1 diabetes, and rheumatoid arthritis, reflecting a shared immunogenetic basis for multiomorbidity burden^44,45^.

**Figure 5.**
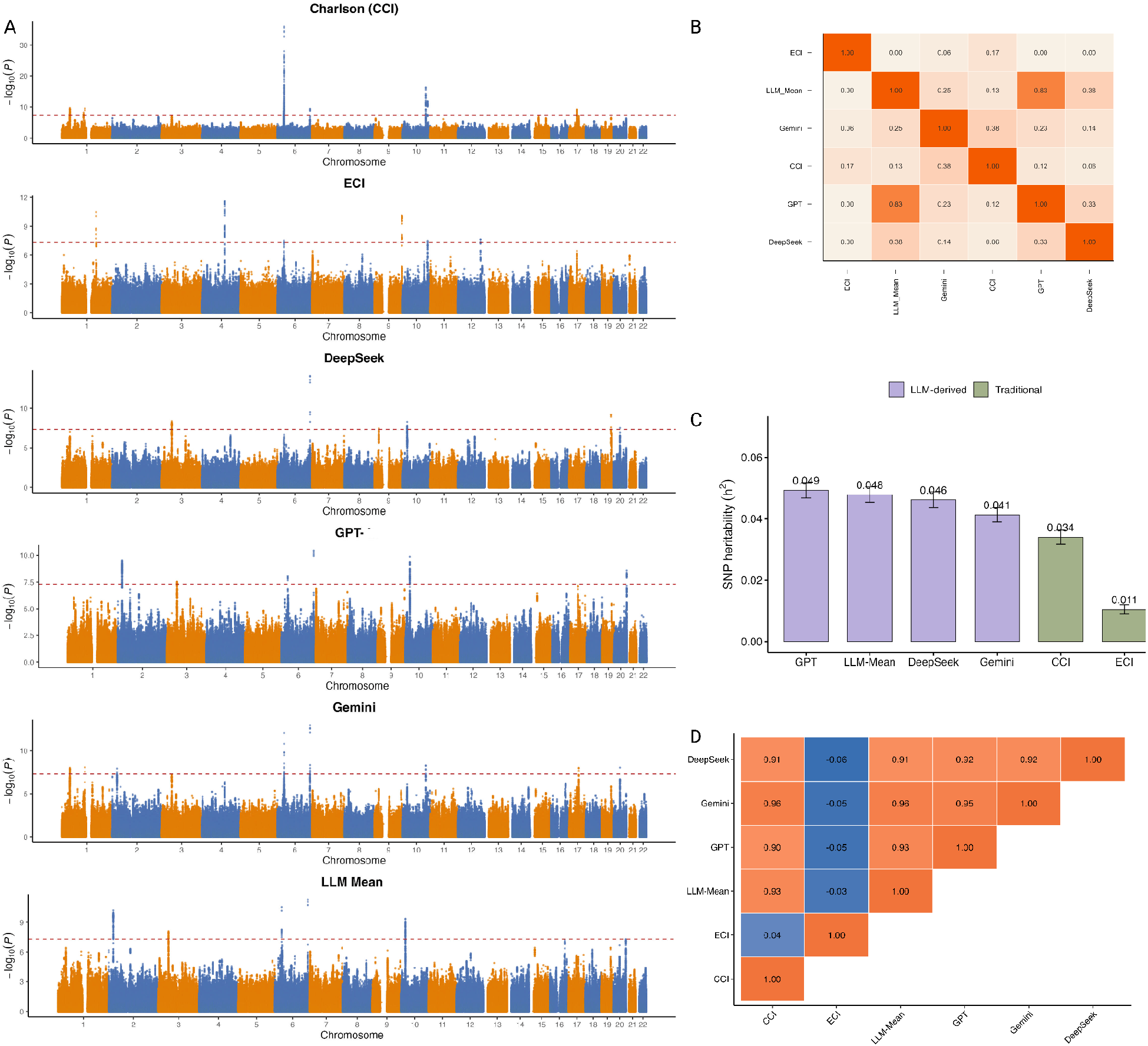
Genome-wide association analysis of CoLLM-derived and traditional multimorbidity traits. (A) Manhattan plots summarizing GWAS results for all four CoLLM-derived phenotypic traits, along with traditional indices CCI andECI. (B) Heatmap showing the Jaccard index of shared significant loci across all traits, reflecting the extent of genetics. For each trait, significant SNPs (P < 5 × 10^-8) were grouped into loci by merging signals within 100 kb on the same chromosome. Pairwise overlap was quantified as the proportion of shared loci relative to the union of loci across the two traits. (C) SNP-based heritability (SNP-h^2^) estimates for each trait. (D) Heatmap of genetic correlations among all traits.

Another consistent and significant association was observed at the *LPA* locus (DeepSeek −log_10_P = 14.07, Gemini = 12.98, LLM-Mean = 11.27, GPT = 10.46, CCI = 9.39). The *LPA* gene encodes lipoprotein(a) [Lp(a)], a well-established mediator of atherosclerosis, thrombosis, and coronary artery disease (CAD). The *LPA* region is strongly associated with cardiovascular diseases due to its role in regulating plasma Lp(a) concentration^46,47^. CoLLM-based models revealed additional loci not observed with traditional indices, highlighting their broader biological scope. Notable examples include *MLLT10* (−log_10_P = 9.87 in LLM-Mean, 9.45 in GPT, and 8.92 in DeepSeek), which encodes a transcriptional regulator frequently involved in chromosomal rearrangements leading to various leukemias, and *AC096570.1* (−log_10_P = 10.24 in LLM-Mean, 9.83 in GPT, and 9.67 in Gemini), a long non-coding RNA predominantly expressed in male germline stem cells^48–50^. SNP-heritability analyses using linkage disequilibrium score regression (LDSC) confirmed heritability and genetic correlation patterns among all traits. Observed SNP-based heritability estimates were h^2^ = 0.034 ± 0.0023 for CCI, 0.046 ± 0.0026 for DeepSeek, 0.041 ± 0.0023 for Gemini, 0.049 ± 0.0025 for GPT-4o, and 0.048 ± 0.0025 for LLM-Mean, indicating that LLM-derived scores capture equal or greater genetic signal than traditional indices **(Fig. 5C)**. LDSC intercepts are 0.985 ± 0.009 (ECI) to 1.064 ± 0.010 (DeepSeek), with values of 1.029 ± 0.010 (CCI), 1.064 ± 0.010 (LLM-Mean), 1.047 ± 0.010 (GPT-4o), and 1.033 ± 0.010 (Gemini), indicating minimal genomic inflation. Interesting patterns were observed for genetic correlations **(Fig. 5D)**. Specifically, genetic correlation analyses revealed strong concordance between CoLLM-derived multimorbidity traits and CCI. The estimated genetic correlations with the CCI were consistently high for all LLM-based models GPT (r_9_ = 0.904, SE = 0.0133), Gemini (r_9_ = 0.9613, SE = 0.0107), DeepSeek (r_9_ = 0.9098, SE = 0.0156), and the LLM-Mean composite (r_9_ = 0.9303, SE = 0.0121). In contrast, when compared with the ECI, the genetic correlations were significantly lower (rho_g ranged from -0.06 to -0.03); this finding is inconsistent with their ∼60% phenotypic correlation (Supplementary Table 5).

This is not surprising because CCI is the sum of the 17 weighted comorbidity categories ^51^. In comparison, ECI is the weighted sum of 30 comorbidities^52^. The observed discrepancy reflects that CCI and ECI capture different biology of comorbidity, and the commodity scores generated by LLMs are more consistent with CCI in both biology and clinical or environmental factors than ECI.

### CoLLM-derived multimorbidity scores exhibit robust associations with disease-specific polygenic risk scores

CoLLM-derived multimorbidity scores showed significant associations with disease-wise polygenic risk across **(Fig. 6).** The LLM-Mean captured the most widespread associations, with cardiometabolic as dominant contributors: type 2 diabetes (β = 0.053, p = 4.4×10^−306^), ischemic stroke (β = 0.054, p = 4.5×10^−331^), hypertension (β = 0.049, p = 1.2×10^−305^), body mass index (BMI; β = 0.044, p = 6.6×10^−285^), coronary artery disease (β = 0.041, p = 3.5×10^−285^), and cardiovascular disease (β = 0.043, p = 2.4×10−248) **(Fig. 6A-B)**. Associations were also observed for immune-mediated conditions like rheumatoid arthritis (β = 0.013, p = 2.7×10^−27^), systemic lupus erythematosus (β = 0.008, p = 6.9×10^−11^), Crohn’s disease (β = 0.008, p = 3.8×10^−12^), and ulcerative colitis (β = 0.007, p = 1.2×10^−9^), reflecting the contribution of systemic inflammation to multimorbidity. Cancer-related PRSs also demonstrated moderate associations, including breast cancer (β = 0.016, p = 5.6×10^−41^), prostate cancer (β = 0.009, p = 1.3×10^−13^), and colorectal cancer (β = 0.009, p = 3.4×10^−13^), consistent with shared hormonal, metabolic, and inflammatory pathways. The Spearman correlation of regression β coefficients between ECI and others was substantially low, which is consistent with the small genetic correlation between ECI and others, confirming the different biology captured by ECI and other scores.

**Figure 6.**
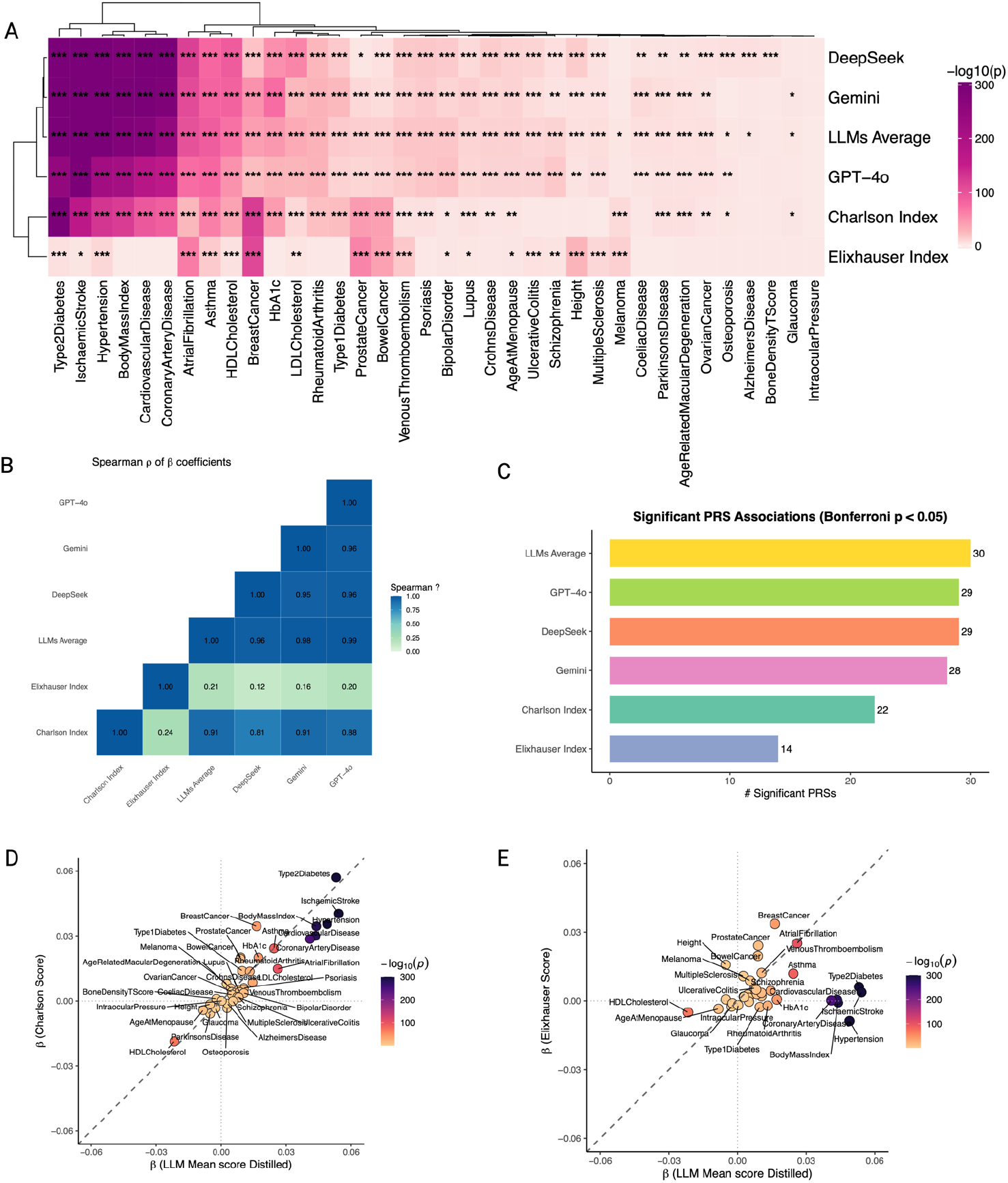
Association between CoLLM-derived multimorbidity scores and polygenic risk scores (PRS). (A) Heatmap showing the statistical significance (*p*-values) of associations derived from linear regression models between CoLLM-derived multimorbidity scores and multiple PRSs across diverse disease domains. Asterisks denote significance thresholds: 1 asterisk (−log_10_ p > 1.3; p < 0.05), 2 asterisks (−log_10_ p > 2; p < 0.01), and 3 asterisks (−log_10_ p > 3; p < 0.001). (B) Spearman correlation of regression β coefficients across models. (C) Bar plot showing the number of significant PRS associations (Bonferroni-adjusted *p* < 0.05) detected for each CoLLM-derived score. (D) Scatterplot illustrating the concordance between CoLLM LLM-mean scores and CCI. (E) Scatterplot illustrating the concordance between CoLLM-mean LLM-derived scores and ECI.

The genetic architecture captured by CoLLM was further highlighted by the number of significant PRS associations after Bonferroni correction. LLM-Mean identified 30 significant traits, exceeding individual models (DeepSeek: 29; GPT-4o: 29; Gemini: 28) and substantially outperforming traditional indices (CCI: 22; ECI: 14) **(Fig. 6C**). Correlation analyses confirmed high cross-model consistency, with LLM-Mean showing uniformly strong agreement with individual LLM-derived scores **(Fig. 6B,D-E**).

### LLM-as-Judge evaluation revealed the meaningfulness of the teacher LLMs and the enhanced capability of the CoLLM student models

Finally, we performed an LLM-as-judge evaluation to qualitatively assess the clinical meaningfulness of Teacher LLMs and compare them with the distilled student CoLLMs **(Supplementary Note 3)**. We used an independent judge Claude Sonnet-4, which is external to all teacher model families to rate each model along three dimensions: (1) Clinical Alignment, (2) Score Reliability, and (3) Outlier & Safety Risk, using a 5-point Likert scale **(See Definition in Supplementary Note 3)**. The LLM-as-a-judge evaluation showed that Teacher LLMs generated generally meaningful multimorbidity scores but exhibited substantial variability and inconsistency across evaluation dimensions **(Supplementary Data 7, Supplementary Figure 2)**. Teacher LLM-Mean was relatively stable across dimensions (clinical alignment: mean = 3.00, median = 3.0; score reliability: mean = 2.97, median = 3.0; safety/outlier risk: mean = 2.90, median = 3.0). However, Teacher Gemini, Teacher GPT-4o, and Teacher DeepSeek showed weaker reliability and/or safety performance (Teacher Gemini reliability: mean = 2.00, median = 2.0; Teacher GPT-4o reliability: mean = 2.10, median = 2.0; Teacher GPT-4o safety/outlier risk: mean = 2.10, median = 2.0; Teacher DeepSeek clinical alignment: mean = 2.00, median = 2.0; Teacher DeepSeek reliability: mean = 1.93, median = 2.0). In contrast, CoLLM student models generally demonstrated improved or more clinically stable behavior relative to their corresponding teacher models. CoLLM-LLM-Mean achieved higher safety/outlier performance (mean = 3.70, median = 4.0), while maintaining stable clinical alignment and reliability (clinical alignment: mean = 3.07, median = 3.0; reliability: mean = 3.13, median = 3.0). CoLLM-GPT-4o improved upon Teacher GPT-4o across reliability and safety/outlier dimensions (reliability: 2.10 to 3.00; safety/outlier risk: 2.10 to 3.07). CoLLM-DeepSeek showed the clearest improvement over Teacher DeepSeek, increasing clinical alignment (2.00 to 3.03), score reliability (1.93 to 3.47), and safety/outlier risk (2.57 to 4.00). Finally, test reliability was strong across repeated fixed-batch evaluations. Across 80 fixed batch-model units and 240 model-level ratings per dimension, Krippendorff’s alpha values were 0.762 for clinical alignment, 0.927 for score reliability, and 0.859 for safety/outlier risk, indicating stable LLM-as-a-judge ratings across repeated runs.

## Discussion

Comorbidities are prevalent and have complex associations with disease severity, mortality, healthcare costs, healthcare utilization, and hospital management. This significance highlights the need for accurate, individual-level, data-driven comorbidity/multimorbidity scoring. Strong multidimensional evaluation should support this effort.

LLMs have made significant advances; their trust in physicians has increased, and their ability to perform complex clinical tasks has achieved expert-level accuracy^53–55^. Despite these promises, deploying LLMs in clinical practice fundamentally requires robust, multidimensional evaluation. This includes not only evaluating traditional clinical tasks such as question answering and text summarization, but also evaluating performance in areas such as genetic contribution and gene set enrichment analysis^56–58^. Strong benchmarks across large and individual-level datasets are necessary to demonstrate LLM reliability, generalizability, and clinical relevance. LLMs offer generative capabilities for new discoveries, yet robust evaluation for complex traits such as comorbidity/multimorbidity remains challenging. Issues include data privacy concerns since models like GPT and Gemini are accessible via APIs with restrictive data privacy policies and the resource-intensive nature of open-source models like Llama-70B. Additionally, large datasets, such as the UK Biobank with over half a million records, make evaluation costly due to the volume of data. Thus, clear evaluation strategies are crucial before integrating LLMs for multimorbidity scoring across diverse healthcare settings. Evaluating LLMs for multimorbidity scores requires efficient, scalable, and privacy-preserving methods.

To address this, we evaluated LLMs using an innovative approach that combined high-fidelity synthetic data with student-model distillation. Since the primary goal of our study was to determine whether LLMs can produce meaningful multimorbidity scores. To answer this question, we applied an independent LLM-as-Judge framework to synthetic data. Through this evaluation, we found that teacher LLM outputs were clinically meaningful but exhibited higher variance. Their meaningfulness and reliability further improved when averaging scores across multiple models rather than using each LLM individually. In contrast, the CoLLM student models were substantially more stable. Given that CoLLM achieves ∼90% concordance with teacher LLMs, its behavior on real UK Biobank data can be interpreted as a privacy-preserved proxy for how teacher LLMs themselves would perform if direct evaluation were permitted. Thus, many of the downstream clinical and genetic insights obtained from CoLLM can reasonably be viewed as reflective of underlying LLMs score generation patterns, while avoiding any exposure of sensitive patient data to external APIs.

CoLLM’s was evaluated for its utility in a large-scale population cohort. We applied all CoLLM variants to electronic health records from the UK Biobank to generate individualized multimorbidity scores and assessed their clinical, genetic, and epidemiological properties. This real-world implementation allowed us to test whether CoLLM-derived scores not only predict clinical outcomes but also align with known biological pathways and heritable disease architecture. Application of CoLLM models to UK Biobank data revealed improvements in mortality risk discrimination compared to traditional multimorbidity indices. In survival analyses, CoLLM-derived scores particularly those from Gemini and the LLM-mean demonstrated superior prognostic discrimination. This enhancement likely reflects CoLLM’s capacity to capture complex, non-additive disease interactions that conventional scoring systems fail to represent. Genome-wide association analyses of CoLLM-derived phenotypes identified multiple biologically plausible loci. Notably, signals at the *HLA-DQ* and *LPA* regions emerged consistently across CoLLM traits, also observed for the CCI. These loci have well-established roles in autoimmune regulation, lipid metabolism, and cardiovascular pathophysiology, suggesting that CoLLM scores reflect true disease biology rather than statistical artifacts. The SNP-based heritability estimates for CoLLM traits exceeded traditional indices, providing further evidence that these metrics capture heritable components of multimorbidity risk. PRS association analyses showed that CoLLM-derived patterns improved association with PRS scores compared to traditional scores.

Although the present study demonstrates the feasibility of the CoLLM framework using UK Biobank data, the generalizability of the findings should be interpreted cautiously. UK Biobank may not fully capture the demographic, clinical, socioeconomic, and health-system diversity present in broader patient populations. Therefore, the performance and calibration of CoLLM may differ when applied to external cohorts with different disease prevalence patterns, coding practices, ancestry composition, or clinical ascertainment processes. In addition, the teacher-student framework in this study relied on a specific set of contemporary LLMs, including GPT-4o, Gemini, and DeepSeek. Although these models were selected to provide diversity across model providers and output behaviors, the resulting teacher scores may not represent the full range of possible LLM score generation patterns. Future work should evaluate the framework using a wider set of teacher models, including open-source, commercial, and biomedical domain-specialized LLMs and should assess the sensitivity of distilled student models to teacher-model selection and prompting strategy. Thus, our results should be interpreted as evidence of feasibility and potential utility within the current study setting rather than as proof of broad applicability across all populations, datasets, or LLM systems. External validation in independent and more diverse cohorts is an essential next step before wider deployment.

In conclusion, our study demonstrates that state-of-the-art large language models (LLMs) GPT-4o, Gemini, and DeepSeek can generate clinically useful information. The developed CoLLMs offer a privacy-preserving clinical tool for multimorbidity risk prediction. A key contribution of this work is the development of a web-based application that enables real-time score generation through an interactive interface, facilitating both validation of CoLLMs and seamless integration into clinical workflows. The application also allows users to download generated scores, supporting further research and downstream analyses of comorbidity and multimorbidity. Overall, this study provides a scalable framework for comorbidity score generation and offers a robust evaluation of its performance across psychiatric, neurological, and oncologic domains, benchmarked against the CCI and ECI indices.

## Limitations

A fundamental limitation of this study is that real-world UK Biobank participant-level data could not be directly provided to closed-source LLMs because of privacy and data-use restrictions. Therefore, teacher LLMs were used only using CTGAN-generated synthetic patient profiles, and LLM performance on real UK Biobank data was inferred indirectly through CoLLM, the distilled student model. Although this design reduces direct exposure of original participant-level records, it does not constitute a formal privacy guarantee, and future work should incorporate locally deployed open-source medical LLMs and formally privacy-preserving synthetic data generation approaches. Second, the fidelity of the synthetic cohort can be improved although the synthetic data showed overall agreement with the real data distribution, we observed discrepancies in the prevalence of specific disease categories, including over-representation of hypertension. Such discrepancies may affect the teacher-student distillation pipeline because over-represented conditions may appear more frequently in synthetic patient profiles and may disproportionately influence teacher LLM scoring. In addition, CTGAN-generated disease co-occurrence patterns may partly reflect artifacts of the synthetic data model rather than true population-level multimorbidity structure, particularly for rare disease combinations and higher-order disease interactions. Third, CoLLM approximates scalar LLM-derived multimorbidity scores rather than transferring full LLM reasoning or explicit clinical reasoning chains. Therefore, the framework should be interpreted as output-level distillation rather than reasoning-level distillation. In addition, teacher-model correlation and LLM-as-Judge scoring primarily measure agreement and consistency rather than clinical correctness, and may inherit biases from the teacher or judge models. Although survival analysis and GWAS provide complementary clinically and biologically grounded evidence, future work should include clinician adjudication, counterfactual patient-profile testing, calibration and error distribution analyses, and validation against external datasets with known clinical outcomes. Finally, all analyses were conducted using a single biobank and a limited set of teacher LLMs, which may limit generalizability across diverse populations, clinical settings, coding systems, and model families. We did not perform detailed cross-population fairness analyses across ethnicity, socioeconomic status, or ancestry groups, nor did we evaluate a broad range of open-source biomedical LLMs because of computational constraints. External validation in more diverse cohorts and systematic evaluation across additional commercial, open-source, and domain-specialized medical LLMs will be important future directions.

## Methods

### Data sources and cohort construction

We used the UK Biobank (UKB), a prospective cohort of ∼500,000 participants recruited between 2006-2010^59^. For this study, we included participants with complete demographic data (age, gender) and diagnostic codes mapped to ICD-10 (UKB study 1712).

For GWAS, we restricted analyses to individuals of European ancestry to minimize population stratification in genetic analyses.

### Representation of diagnoses

To stabilize sparsity and reduce redundancy, ICD-10 codes were aggregated into 263 clinically coherent “parent” categories using a hierarchical grouping scheme derived from (UKB Data-Coding 19)^60,61^ (**Supplementary data 1)**. For each participant (or synthetic record), the diagnosis vector was encoded as binary presence/absence across these categories, alongside age (years) and gender(binary: female/male, ethnicity as additional features. Instead of considering diseases at the individual ICD-10 code level, we used grouped disease categories to reduce data sparsity and token size in the LLM input, which also helped lower the LLM-related experimental costs.

### Synthetic data generation

Because UKB raw data cannot be sent to external APIs under the data-use agreement, we generated synthetic tabular cohorts using CTGAN^37^. A training set of 50,000 and a held-out test set of 10,000 synthetic records were generated from the empirical joint distribution of the real cohort’s grouped ICD-10 categories, age, and sex. CTGAN hyperparameters were set to [epochs =100, batch size = 5000]. The generated synthetic data were evaluated using the learning the disease prevalence between real and synthetic datasets^62^. To assess comorbidity patterns, the Jaccard index between pairs of diseases was calculated for both real and synthetic data, followed by analysis of the Pearson correlation between them. We also evaluated the disclosure protection of synthetic data which measures the risk associated with disclosing the synthetic data. We used SDmetrics from the Synthetic Data Vault (SDV) evaluation framework to perform disclosure protection^41^. This analysis provides an empirical estimate of attribute-disclosure risk and helps assess whether the synthetic data may reveal sensitive disease-pattern information. The synthetic cohort was therefore used to capture aggregate ICD-10 disease-pattern structure while reducing direct exposure of original participant-level records and identifiers during downstream model evaluation. However, this metric should be interpreted as a heuristic privacy-risk assessment rather than a formal privacy guarantee. The CTGAN model was not trained with differential privacy, and formal privacy audits such as membership-inference or attribute-inference attacks were not performed in this study.

### Teacher large language models and scoring protocol

We utilized three state-of-the-art large language models, GPT-4o, Gemini, and DeepSeek, to generate comorbidity scores using a zero-shot prompt on synthetic data. The temperature value was set to 0 to ensure that the models produced more deterministic responses.

### Student model (CoLLM) architecture and training

Next, we trained student models, referred to as CoLLM models to mimic the multimorbidity score predictions generated by the teacher LLMs. Each CoLLM model was implemented as a multilayer perceptron (MLP) regression network trained on structured patient-level features derived from the synthetic dataset. The full synthetic dataset consisted of 60,000 samples, of which 50,000 were used for training and 10,000 were held out for testing to evaluate how closely the student models reproduced the teacher LLM-derived scores. The input features included demographic variables and ICD-10-derived disease indicators. Categorical variables were one-hot encoded, and the held-out test set was aligned to the same feature columns as the training set. The target variables were the multimorbidity scores generated by the teacher LLMs. For each teacher LLM target, including GPT-4o, Gemini, and DeepSeek, a separate CoLLM student model was trained. In addition, a fourth student model was trained using the mean multimorbidity score across the three teacher LLMs as the target.

For each target, the MLP architecture was optimized using up to 25 hyperparameter trials. The hyperparameter search varied network depth, hidden layer width, dropout rate, activation function, learning rate, weight decay, and batch size. The search space included 3–6 hidden layers, hidden layer widths selected from 128, 256, 512, and 1024 units, ReLU or LeakyReLU activation functions, dropout values ranging from 0.1 to 0.4, learning rates of 1e-4, 3e-4, and 1e-3, weight decay values of 0, 1e-6, 1e-5, and 1e-4, and batch sizes of 64, 128, 256, and 512. For hyperparameter selection, 10% of the training data was used as a validation set. Models were trained for a maximum of 200 epochs using mean squared error loss and the Adam optimizer. Early stopping was applied based on validation loss with a patience of 12 epochs. A ReduceLROnPlateau learning-rate scheduler was used to reduce the learning rate when validation loss plateaued, and gradient clipping with a maximum norm of 1.0 was applied during training. The optimal configuration for each target was selected based on the lowest validation loss. The best-performing architecture was then retrained on the full training set and evaluated on the held-out test set.

### Evaluation of CoLLMs

We evaluated the CoLLMs on 10,000 test samples by computing the Spearman correlation and R^2^ between the true LLM scores (teacher model scores) and the CoLLM-predicted scores. We also assessed model calibration by binning the true scores into five quintiles and computing the mean predicted versus mean true score within each bin for each model. Calibration curves were generated by plotting these bin-wise averages against the identity line to visualize systematic over- or under-prediction.

### LLMs-as-a-Judge Evaluation

Using LLMs as a judge is a recent method for evaluating the output of other LLMs, a practice that is particularly useful when ground truth information is unavailable. LLMs-as-a-judge present a strong alternative to traditional expert-driven qualitative evaluations^63–65^.

In our study, we used this method to evaluate both Teacher LLMs and CoLLM scores (**Supplementary Figure 1)**. We used the Claude-Sonnet model from the Anthropic LLM family as the independent judge. This model is completely separate from the multimorbidity-scoring models, thereby avoiding kinship bias that could arise when a model evaluates another from the same family^66^.

The evaluation was conducted on a synthetic test dataset of 10,000 samples. The judge model was given a zero-shot instruction prompt **(see Supplementary Note 3**) and configured with a temperature of 0. To ensure reproducibility and assess the stability of the LLM-as-judge rankings, we performed the evaluation using a fixed-batch test-retest design. Specifically, we constructed 10 fixed patient batches each containing 100 patient records sampled from the generated test data. Each fixed batch was evaluated three times using the same LLM-as-judge prompt and judge model. This design resulted in 30 repeated evaluations per model for each scoring dimension. The judge’s task was to rate all multimorbidity scoring models on a 1-5 Likert scale across three dimensions: 1)Clinical Alignment 2)Score Reliability 3)Outlier and Safety Risk **(Detailed definitions for each dimension can be found in the Supplementary Note)**. The judge model analyzed ICD-10 code patterns and demographic information to assign its ratings.

### Scoring the real UK Biobank cohort

Finally, using the student models, we predicted the comorbidity scores on the real UK Biobank data.

### Evaluation

We performed survival analysis, genome-wide association studies (GWAS), and correlation analyses between comorbidity scores and polygenic risk scores (PRS) to evaluate the LLM-generated comorbidity scores and benchmark them against traditional comorbidity indices.

### Survival analyses

We performed survival analysis to evaluate the association between the predicted comorbidity scores and overall mortality in the UK Biobank cohort. The event of interest was death. Participants without a recorded death date were censored on 2022-11-30. The time-to-event variable was defined as the duration (in days) from the date of score calculation (the last diagnosis date) to the date of death or censoring. Participants’s age, sex, and ethnicity were included as covariates. We fitted Cox proportional hazards regression models to assess the association between the predicted comorbidity scores and all-cause mortality. Model performance was evaluated using the concordance index (C-index) to measure predictive discrimination. All analyses were performed age-group–wise, given the strong influence of age on survival outcomes. We stratified participants into five categories based on their reference age at the time of score calculation: <50 years, 50–59 years, 60–69 years, 70–79 years, and ≥80 years. Time-dependent AUC was calculated at 1, 3, and 5 years to evaluate mortality risk discrimination at clinically relevant follow-up intervals, complementing the overall Cox model C-index.

### Genome-wide association studies (GWAS)

Genome-wide association analyses were performed using the GRAB framework, which efficiently accounts for population structure and cryptic relatedness in large-scale biobank data^42^. We first constructed a sparse genetic relatedness matrix (sGRM) from UK Biobank genotype data. For association analysis chromosome-specific datasets were prepared with a filter (MAF ≥ 0.01). Phenotypes of interest including 4-level ordinal groupings of Charlson and LLM-derived comorbidity scores were analyzed with age, sex, and the top 10 principal components as covariates. Null models were fit with GRAB using the proportional odds logistic mixed model (POLMM) for ordinal traits, with a maximum likelihood fallback if convergence failed; hyperparameters included tolerance thresholds for variance components (tolTau = 0.2) and fixed effects (tolBeta = 0.1). Single-variant association testing was then performed for each chromosome, with results combined across the genome. Statistical significance was defined at the conventional genome-wide threshold (P < 5×10^−8^). Quality control included evaluation of test statistic inflation (λGC), quantile–quantile plots, and Manhattan plots. All analyses were conducted on a high-performance computing cluster with PLINK2 for preprocessing and GRAB (R) for sGRM and association test. Genetic correlations and heritability estimation were performed using the web-based platform Complex-Traits Genetics Virtual Lab (CTG-VL)^67^.

### Association of the Comorbidity score and the Polygenic Risk Score

We also assessed the association between the comorbidity scores and polygenic risk scores (PRS). Disease-specific PRS data were obtained from the UK Biobank data field 26202. We then performed linear regression analyses to evaluate these associations, reporting both the β coefficients and corresponding p-values to quantify effect size and statistical significance.

### Model Deployment

Finally, we have also provided an open-source web app for real-time comorbidity score prediction using CoLLM (https://zhulab-collm.streamlit.app/). This interface enables users to input ICD-10 code lists and demographic information to obtain CoLLM-derived comorbidity scores in real time, supporting transparency and external validation. Additional details for using instructions of our app are provided in the **Supplementary Note 1**.

## Data Availability

All secondary data are available at https://github.com/rxa615/CoLLM, and the CoLLM web application is accessible at https://zhulab-collm.streamlit.app/. Primary data can be obtained from the UK Biobank.

## Acknowledgments

We would like to acknowledge the Ohio Supercomputer Center for providing computational resources.

## Author Contribution

Study Design: XZ, RA

Data and Model Selection: XZ, RA

Analysis: RA, YY, ML

Paper Writing: XZ, RA

## References

1. Chowdhury, S. R., Chandra Das, D., Sunna, T. C., Beyene, J. & Hossain, A. Global and regional prevalence of multimorbidity in the adult population in community settings: a systematic review and meta-analysis. EClinicalMedicine 57, 101860 (2023).

2. Haug, N. et al. High-risk multimorbidity patterns on the road to cardiovascular mortality. BMC Medicine 18, 1–12 (2020).

3. Skou, S. T. et al. Multimorbidity. Nature reviews. Disease primers 8, 48 (2022).

4. Johnston, M. C., Crilly, M., Black, C., Prescott, G. J. & Mercer, S. W. Defining and measuring multimorbidity: a systematic review of systematic reviews. Eur J Public Health 29, 182–189 (2019).

5. Valderas, J. M., Starfield, B., Sibbald, B., Salisbury, C. & Roland, M. Defining Comorbidity: Implications for Understanding Health and Health Services. Annals of Family Medicine 7, 357 (2009).

6. Zhao, X. et al. Associations of multimorbidity with mortality, hospital stay, and hospitalization costs in Chinese surgical patients: a retrospective cohort study. BMC Anesthesiology 25, 407 (2025).

7. Dervić, E. et al. Unraveling cradle-to-grave disease trajectories from multilayer comorbidity networks. npj Digital Medicine 7, 1–12 (2024).

8. König, S., Vaskyte, U., Boesing, M., Lüthi-Corridori, G. & Leuppi, J. D. The Role of Comorbidities in COVID-19 Severity. Viruses 17, 957 (2025).

9. Nigatu, B. Z. & Dessie, N. T. Prevalence of comorbidities and their association with disease severity and mortality in COVID-19 patients: A systematic review and meta-analysis. Journal of multimorbidity and comorbidity 15, (2025).

10. Zhang, J. et al. Comorbidity patterns associated with severe COVID-19 outcomes: A cohort study based on the UK Biobank. PLOS ONE 20, e0329701 (2025).

11. Rashid, M. et al. Impact of co-morbid burden on mortality in patients with coronary heart disease, heart failure, and cerebrovascular accident: a systematic review and meta-analysis. European heart journal. Quality of care & clinical outcomes 3, (2017).

12. Cruz-Ávila, H. A., Ramírez-Alatriste, F., Martínez-García, M. & Hernández-Lemus, E. Comorbidity patterns in cardiovascular diseases: the role of life-stage and socioeconomic status. Front. Cardiovasc. Med. 11, 1215458 (2024).

13. Søgaard, M., Thomsen, R. W., Bossen, K. S., Sørensen, H. T. & Nørgaard, M. The impact of comorbidity on cancer survival: a review. Clinical epidemiology 5, (2013).

14. George, M., Smith, A., Sabesan, S. & Ranmuthugala, G. Physical Comorbidities and Their Relationship with Cancer Treatment and Its Outcomes in Older Adult Populations: Systematic Review. JMIR Cancer 7, e26425 (2021).

15. Comparison of established comorbidity scores using administrative data of patients undergoing surgery or interventional procedures in Massachusetts. Journal of Clinical Epidemiology 185, 111869 (2025).

16. Sharma, N., Schwendimann, R., Endrich, O., Ausserhofer, D. & Simon, M. Comparing Charlson and Elixhauser comorbidity indices with different weightings to predict in-hospital mortality: an analysis of national inpatient data. BMC health services research 21, (2021).

17. Baron, R. B. et al. A Comparison of the Elixhauser and Charlson Comorbidity Indices: Predicting In-Hospital Complications Following Anterior Lumbar Interbody Fusions. World Neurosurg 144, e353–e360 (2020).

18. Brämer, G. R. International statistical classification of diseases and related health problems. Tenth revision. World health statistics quarterly. Rapport trimestriel de statistiques sanitaires mondiales 41, (1988).

19. Quer, G. & Topol, E. J. The potential for large language models to transform cardiovascular medicine. Lancet Digit Health 6, e767–e771 (2024).

20. Lin, C. & Kuo, C.-F. Roles and potential of Large language models in healthcare: A comprehensive review. Biomedical Journal 48, 100868 (2025).

21. Singhal, K. et al. Large language models encode clinical knowledge. Nature 620, 172–180 (2023).

22. Qiu, J. et al. LLM-based agentic systems in medicine and healthcare. Nature Machine Intelligence 6, 1418–1420 (2024).

23. Kung, T. H. et al. Performance of ChatGPT on USMLE: Potential for AI-assisted medical education using large language models. PLOS Digital Health 2, e0000198 (2023).

24. Gaber, F. et al. Evaluating large language model workflows in clinical decision support for triage and referral and diagnosis. NPJ Digital Medicine 8, 263 (2025).

25. Singhal, K. et al. Toward expert-level medical question answering with large language models. Nature Medicine 31, 943–950 (2025).

26. Meskó, B. & Topol, E. J. The imperative for regulatory oversight of large language models (or generative AI) in healthcare. NPJ Digit Med 6, 120 (2023).

27. Wiest, I. C. et al. Privacy-preserving large language models for structured medical information retrieval. npj Digital Medicine 7, 257 (2024).

28. Zhong, X. et al. Considerations for Patient Privacy of Large Language Models in Health Care: Scoping Review. Journal of Medical Internet Research 27, e76571 (2025).

29. Website. UK Biobank. (2023, October 26). Material transfer agreement: New applicant MTA – Data only (ACCESS_031_F v1.4). UK Biobank Limited. https://www.ukbiobank.ac.uk/wp-content/uploads/2025/01/Material-Transfer-Agreement.pdf.

30. Moëll, B., Sand Aronsson, F. & Akbar, S. Medical reasoning in LLMs: an in-depth analysis of DeepSeek R1. Front Artif Intell 8, 1616145 (2025).

31. Bedi, S., Jiang, Y., Chung, P., Koyejo, S. & Shah, N. Fidelity of Medical Reasoning in Large Language Models. JAMA Netw Open 8, e2526021 (2025).

32. Shojaee, P., et al. The Illusion of Thinking: Understanding the Strengths and Limitations of Reasoning Models via the Lens of Problem Complexity. (2025).

33. Mirzadeh, I., et al. GSM-Symbolic: Understanding the Limitations of Mathematical Reasoning in Large Language Models. (2024).

34. Hinton, G., Vinyals, O. & Dean, J. Distilling the Knowledge in a Neural Network. (2015).

35. Iyer, S. When Babies Teach Babies: Can student knowledge sharing outperform Teacher-Guided Distillation on small datasets? in The 2nd BabyLM Challenge at the 28th Conference on Computational Natural Language Learning 197–211 (2024).

36. Can Students Beyond The Teacher? Distilling Knowledge from Teacher’s Bias. https://arxiv.org/html/2412.09874v1.

37. Xu, L., Skoularidou, M., Cuesta-Infante, A. & Veeramachaneni, K. Modeling Tabular data using Conditional GAN. Advances in Neural Information Processing Systems 32, (2019).

38. OpenAI et al. GPT-4 Technical Report. (2023).

39. Anil, R. et al. Gemini: A Family of Highly Capable Multimodal Models. (2023).

40. Guo, D. et al. DeepSeek-R1 incentivizes reasoning in LLMs through reinforcement learning. Nature 645, 633–638 (2025).

41. Patki, N., Wedge, R. & Veeramachaneni, K. The synthetic data vault. in 2016 IEEE International Conference on Data Science and Advanced Analytics (DSAA) (IEEE, 2016). doi:10.1109/dsaa.2016.49.

42. Bi, W. et al. Efficient mixed model approach for large-scale genome-wide association studies of ordinal categorical phenotypes. American journal of human genetics 108, (2021).

43. Genome-Wide Robust Analysis for Biobank Data (GRAB) [R package GRAB version 0.2.3]. (2025).

44. Gough, S. C. L. & Simmonds, M. J. The HLA Region and Autoimmune Disease: Associations and Mechanisms of Action. Current Genomics 8, 453 (2007).

45. van Belzen, M. J. et al. Defining the contribution of the HLA region to cis DQ2-positive coeliac disease patients. Genes Immun 5, 215–220 (2004).

46. Clarke, R. et al. Genetic Variants Associated with Lp(a) Lipoprotein Level and Coronary Disease. (2009) doi:10.1056/NEJMoa0902604.

47. El-Menyar, A., Khan, N. A., Al Mahmeed, W., Al Suwaidi, J. & Al-Thani, H. Cardiovascular Implications of Lipoprotein(a) and its Genetic Variants: A Critical Review From the Middle East. JACC: Asia (2025) doi:10.1016/j.jacasi.2025.04.012.

48. Uğurlu-Çimen, D. et al. AF10 (MLLT10) prevents somatic cell reprogramming through regulation of DOT1L-mediated H3K79 methylation. Epigenetics & chromatin 14, (2021).

49. Deutsch, J. L. & Heath, J. L. MLLT10 in benign and malignant hematopoiesis. Experimental hematology 87, (2020).

50. Jing, X. et al. MLLT10 promotes tumor migration, invasion, and metastasis in human colorectal cancer. Scandinavian journal of gastroenterology 53, (2018).

51. Ghali, W. A., Hall, R. E., Rosen, A. K., Ash, A. S. & Moskowitz, M. A. Searching for an improved clinical comorbidity index for use with ICD-9-CM administrative data. J Clin Epidemiol 49, 273–278 (1996).

52. van Walraven, C., Austin, P. C., Jennings, A., Quan, H. & Forster, A. J. A modification of the Elixhauser comorbidity measures into a point system for hospital death using administrative data. Med Care 47, 626–633 (2009).

53. Alohali, K. I., Almusaeeb, L. A., Almubarak, A. A., Alohali, A. I. & Muaygil, R. A. Reasoning-based LLMs surpass average human performance on medical social skills. Scientific Reports 15, 1–9 (2025).

54. Luo, X. et al. Large language models surpass human experts in predicting neuroscience results. Nature Human Behaviour 9, 305–315 (2024).

55. LLMs model how humans induce logically structured rules. Journal of Memory and Language 146, 104675 (2026).

56. Wang, Z. et al. GeneAgent: self-verification language agent for gene-set analysis using domain databases. Nature Methods 22, 1677–1685 (2025).

57. Enhancing functional gene set analysis with large language models. Nature Methods 22, 22–23 (2024).

58. Zhu, J., et al. Enhancing gene set overrepresentation analysis with large language models. Bioinform Adv 5, vbaf054 (2025).

59. Sudlow, C. et al. UK Biobank: An Open Access Resource for Identifying the Causes of a Wide Range of Complex Diseases of Middle and Old Age. PLOS Medicine 12, e1001779 (2015).

60. Watzlaf, V. J. M., Garvin, J. H., Moeini, S. & Anania-Firouzan, P. The Effectiveness of ICD-10-CM in Capturing Public Health Diseases. Perspectives in Health Information Management / AHIMA, American Health Information Management Association 4, 6 (2007).

61. Quan, H. et al. Coding algorithms for defining comorbidities in ICD-9-CM and ICD-10 administrative data. Medical care 43, (2005).

62. Cui, E. H., Li, Y. & Liu, Z. The Kolmogorov-Smirnov Statistic Revisited. (2025).

63. Croxford, E. et al. Evaluating clinical AI summaries with large language models as judges. npj Digital Medicine 8, 640 (2025).

64. Williams, G., Rutunda, S., Nzabakira, F. & Mateen, B. A. Human Evaluators vs. LLM-as-a-Judge: Toward Scalable, Real-Time Evaluation of GenAI in Global Health. medRxiv 2025.10.27.25338910 (2025) doi:10.1101/2025.10.27.25338910.

65. Li, H., et al. LLMs-as-Judges: A Comprehensive Survey on LLM-based Evaluation Methods. (2024).

66. Walker, R. S. & Hill, K. R. Causes, Consequences, and Kin Bias of Human Group Fissions. Human Nature 25, 465–475 (2014).

67. Website. https://www.biorxiv.org/content/10.1101/518027v4.

65. Cuellar-Partida, Gabriel, et al. "Complex-Traits Genetics Virtual Lab: A community-driven web platform for post-GWAS analyses." BioRxiv (2019): 518027.

