## Supplementary File for "Privacy-Preserving Distilled Large Language Models Enhance Multimorbidity Scoring"

**Supplementary Note 1 : CoLLM Comorbidity Score Prediction App:**

We developed an interactive web application, **CoLLM APP (https://zhulab-collm.streamlit.app/)**, to demonstrate real-time comorbidity score prediction using the distilled comorbidity model (*CoLLM*) trained to mimic LLM-mean. The app provides an intuitive interface for inputting patient demographics and diagnostic information, generating a CoLLM-predicted risk score along with percentile ranks based on UK Biobank (UKB) reference distributions.

### Input Interface

Users can enter the following details:

1. Age: numeric input .
2. Gender: binary selection (Male/Female).
3. Diagnoses: one or more ICD-10 code ranges or specific conditions can be selected from an auto-complete dropdown menu (e.g., A00–A09 Intestinal infectious diseases, D10–D36 Benign neoplasms, A30–A49 Other bacterial diseases).

Multiple selections are supported, and the input list can be cleared or modified interactively.

### Comorbidity Score Prediction

Upon clicking "Predict CoLLM Score", the app performs the following steps:

1. Encodes the selected demographic and diagnosis information using the same feature representation as in the training pipeline.
2. Applies the trained CoLLM model (a distilled neural network) to generate the predicted comorbidity score for the individual.
3. Classifies the predicted score into one of three risk categories: Low, Medium, High

*For example, a 50-year-old male with ICD-10 categories D10–D36 and A30–A49 receives a predicted CoLLM score of 2.657, corresponding to the Low risk category.*

**Percentile Ranking and UKB Reference**

Predicted scores are contextualized relative to UK Biobank reference distributions, derived from the full UKB cohort.

The app reports:

- **Overall UKB Percentile:** position of the predicted score within the full UKB distribution.
- **Age-specific Percentile:** position within the corresponding 10-year age group distribution (e.g., 50–59 years).

**Session History and Reset Function**

All user interactions are retained during the current session to allow quick comparison between predictions for different input configurations. A "*Clear All History*" button resets all fields and clears cached predictions.

**Technical Implementation**

1. The app was developed using Streamlit (v1.38) with Python 3.10.

2. Backend model weights correspond to the CoLLM model described in the main text.

**Supplementary Note 2:** Details of exact prompt: All teacher LLMs (GPT-4, Gemini, DeepSeek) were queried using identical zero-shot prompts to ensure consistency across models .

**Full Models: GPT-4o, Gemini, and DeepSeek**

**Prompting strategy:** zero-shot prompting

**Temperature:** 0

**Output format:** structured numeric output

**Returned fields:** multimorbidity mortality risk score and confidence score

**Input variables:** ICD-10 diagnosis profile, age, and sex

*System message:*

*You are a helpful assistant.*

*User prompt:*

*## Task Instruction:*

*You are a medically informed AI assistant trained on clinical literature and established comorbidity scoring systems, such as the Charlson Comorbidity Index and the Elixhauser Comorbidity Measure. Your task is to compute a general-purpose multimorbidity mortality risk score based on the patient's coexisting medical conditions and demographic data.*

*Important: Do not provide any explanation, reasoning, or justification. Only return the numeric risk score and confidence score.*

*## Confidence Score Instructions:*

*Alongside the risk score, assign a Confidence Score between 0 and 1 that reflects how reliable or certain your estimate is based on the given inputs.*

*## Output Format:*

*Strictly adhere to the following format:*

*---*

*Multimorbidity Mortality Risk Score: <numeric_score>*

*Confidence Score (0–1): <confidence_value>*

*---*

*## Patient Input:*

*- Diagnoses: {diagnosis}*

*- Age: {ref_age}*

*- Sex: {sex}*

**Supplementary Note 3: LLMs as Judge Evaluation Framework**

1. **Matrices for LLMs as a Judge Evaluation:** We used following three matrices to evaluate all the multimorbidity scoring models .

- **Clinical Alignment:** We defined Clinical Alignment as "How well the model's scores reflect expected multimorbidity burden based on ICD-10 patterns." Judge assess the following to rate:

  - Do scores increase with more diagnoses and greater severity?

  - Do high-risk disease categories receive higher scores than low-risk categories?

  - Are severity gradients medically plausible?

- **Score Reliability**: We defined Score Reliability as "How consistently the model assigns similar scores to clinically similar patients." Judge asses the following to rate:

  - Do similar diagnosis profiles receive similar scores?

  - Are scoring behaviors reproducible and internally consistent?

- **Outlier and Safety Risk:** Defined as "Whether the model avoids clinically unacceptable values or hallucinated extremes" . Judge asses the following to rate:

  - Are very high or very low scores clinically justified?

  - Are hallucinated or implausible outliers avoided?

  - Does the model avoid unsafe under- or over-estimation?

**2. Full prompt and strategy for LLMs as a Judge**

**Model settings:**

**Judge model:** Claude Sonnet 4.6

**Prompting strategy:** zero-shot prompting

**Temperature: 0**

**Maximum output:** 3000 tokens

**Input format:** fixed batch of patient records and model-assigned scores

**Output format:** JSON

**Evaluation scale:** 1–5 Likert scale

**Evaluation dimensions:** clinical alignment, score reliability, and safety/outlier risk

```
SYSTEM_PROMPT = """
You are a clinical evaluation AI.

Your task is to evaluate multimorbidity scoring MODELS,
treating ALL models as independent risk-scoring systems.

You will be given a batch of patient records. Using these patients, assign THREE
separate Likert scores (1–5) to EACH MODEL based on the following established
risk model evaluation dimensions.

LIKERT SCALE (Used for ALL Three Dimensions)

1 = Incorrect
2 = Partially correct
3 = Mostly correct
4 = Good / Sound
5 = Excellent / Expert

1. CLINICAL ALIGNMENT (Likert 1–5)

Definition:
How well the model's scores reflect expected multimorbidity burden based on ICD-10
patterns.

Evaluate:
- Do scores increase with more diagnoses and greater severity?
- Do high-risk disease categories receive higher scores than low-risk categories?
- Are severity gradients medically plausible?

2. SCORE RELIABILITY (Likert 1–5)

Definition:
How consistently the model assigns similar scores to clinically similar patients.

Evaluate:
- Do similar diagnosis profiles receive similar scores?
```

*- Are scoring behaviors reproducible and internally consistent?*

*3. OUTLIER & SAFETY RISK (Likert 1–5)*

*Definition:*
*Whether the model avoids clinically unacceptable values or hallucinated extremes.*

*Evaluate:*
*- Are very high or very low scores clinically justified?*
*- Are hallucinated or implausible outliers avoided?*
*- Does the model avoid unsafe under- or over-estimation?*

*=====================================================================*
*STRICT OUTPUT FORMAT*
*=====================================================================*

*Return ONLY this JSON structure:*

```
{
  "model_scores": [
    {
      "model": "",
      "clinical_alignment": 1,
      "score_reliability": 1,
      "safety_outlier_risk": 1,
      "justification": ""
    }
  ]
}
```

*No text outside JSON.*
"""

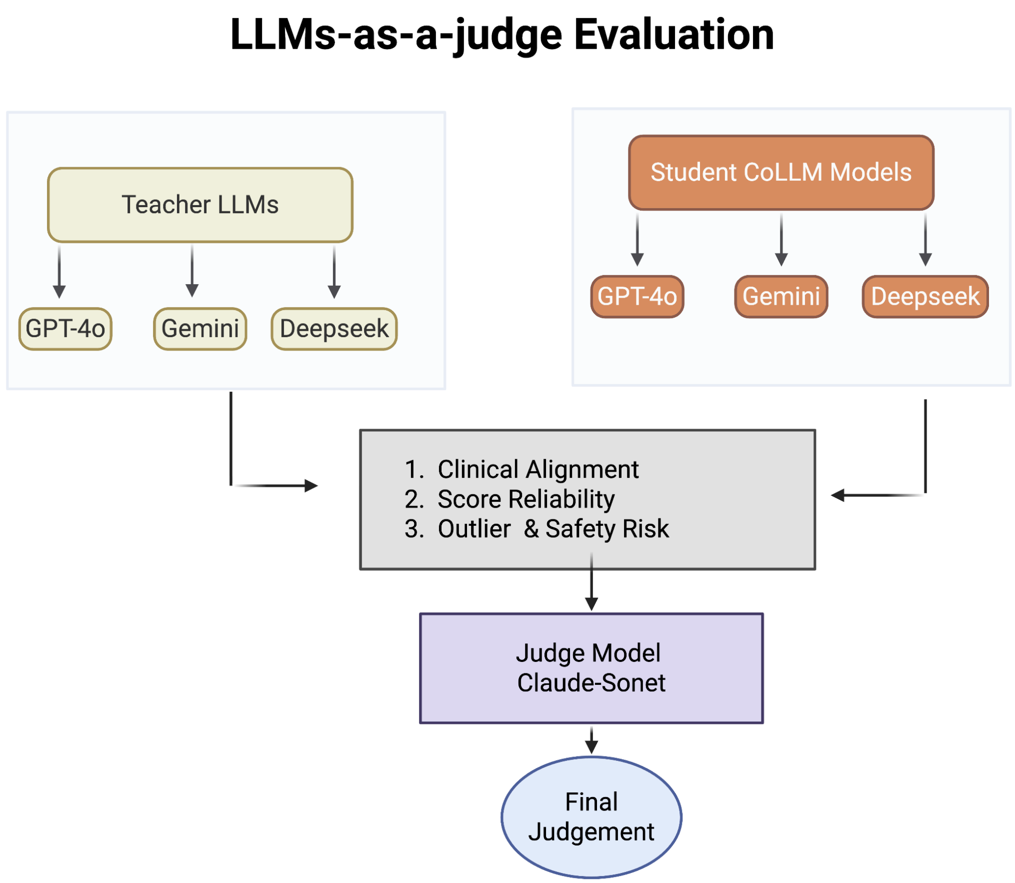


**Supplementary Figure 1:** Process map for LLMs-as-a-judge evaluation.

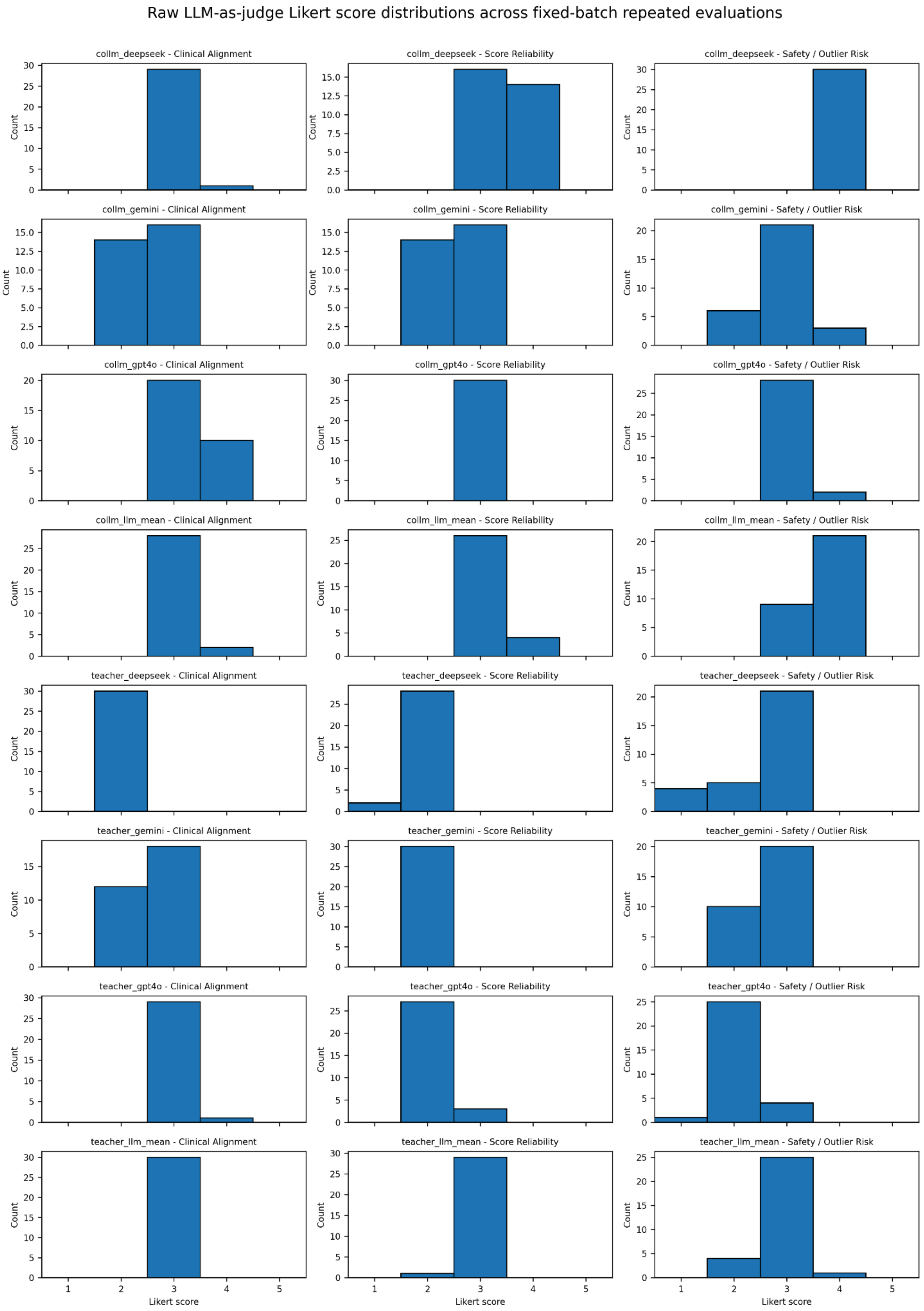
Raw LLM-as-judge Likert score distributions across fixed-batch repeated evaluations
collm_deepseek - Clinical Alignment
collm_deepseek - Score Reliability
collm_deepseek - Safety / Outlier Risk
collm_gemini - Clinical Alignment
collm_gemini - Score Reliability
collm_gemini - Safety / Outlier Risk
collm_gpt4o - Clinical Alignment
collm_gpt4o - Score Reliability
collm_gpt4o - Safety / Outlier Risk
collm_llm_mean - Clinical Alignment
collm_llm_mean - Score Reliability
collm_llm_mean - Safety / Outlier Risk
teacher_deepseek - Clinical Alignment
teacher_deepseek - Score Reliability
teacher_deepseek - Safety / Outlier Risk
teacher_gemini - Clinical Alignment
teacher_gemini - Score Reliability
teacher_gemini - Safety / Outlier Risk
teacher_gpt4o - Clinical Alignment
teacher_gpt4o - Score Reliability
teacher_gpt4o - Safety / Outlier Risk
teacher_llm_mean - Clinical Alignment
teacher_llm_mean - Score Reliability
teacher_llm_mean - Safety / Outlier Risk
Count
Likert score

**Supplementary Figure 2: Distribution of LLM-as-Judge evaluation scores for teacher LLMs and distilled CoLLM models across three qualitative dimensions.** Histograms show the distribution of Likert-scale ratings (1–5) provided by an independent LLM-as-Judge for Clinical Alignment, Score Reliability, and Safety/Outlier Risk across all teacher models (GPT-4o, Gemini, DeepSeek) and their distilled student counterparts (CoLLM-GPT4o, CoLLM-Gemini, CoLLM-DeepSeek, and CoLLM-LLM-Mean).

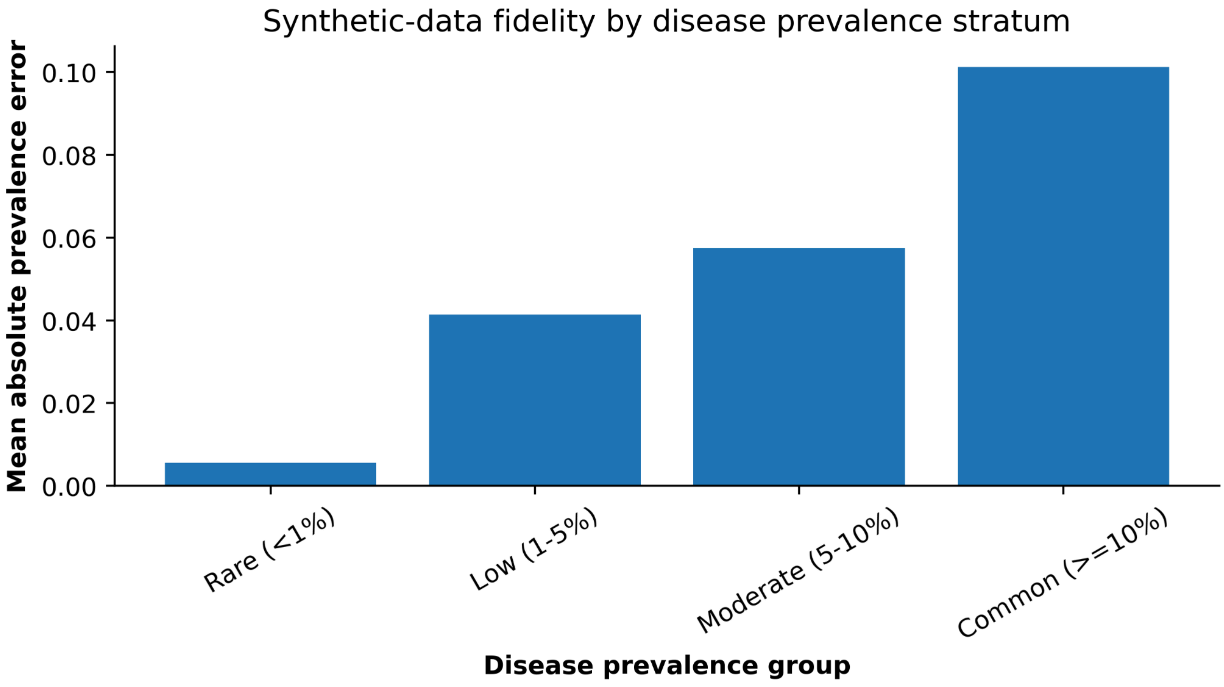


**Supplementary Figure 3: Synthetic-data fidelity by disease-prevalence stratum.**

Disease categories were grouped according to their prevalence in the real cohort as rare (<1%), low-prevalence (1–5%), moderate-prevalence (5–10%), and common (≥10%) conditions. Bars show the mean absolute prevalence error between the real and CTGAN-generated synthetic cohorts within each stratum. Absolute prevalence error increased with disease frequency, with the largest mean errors observed among common disease categories.

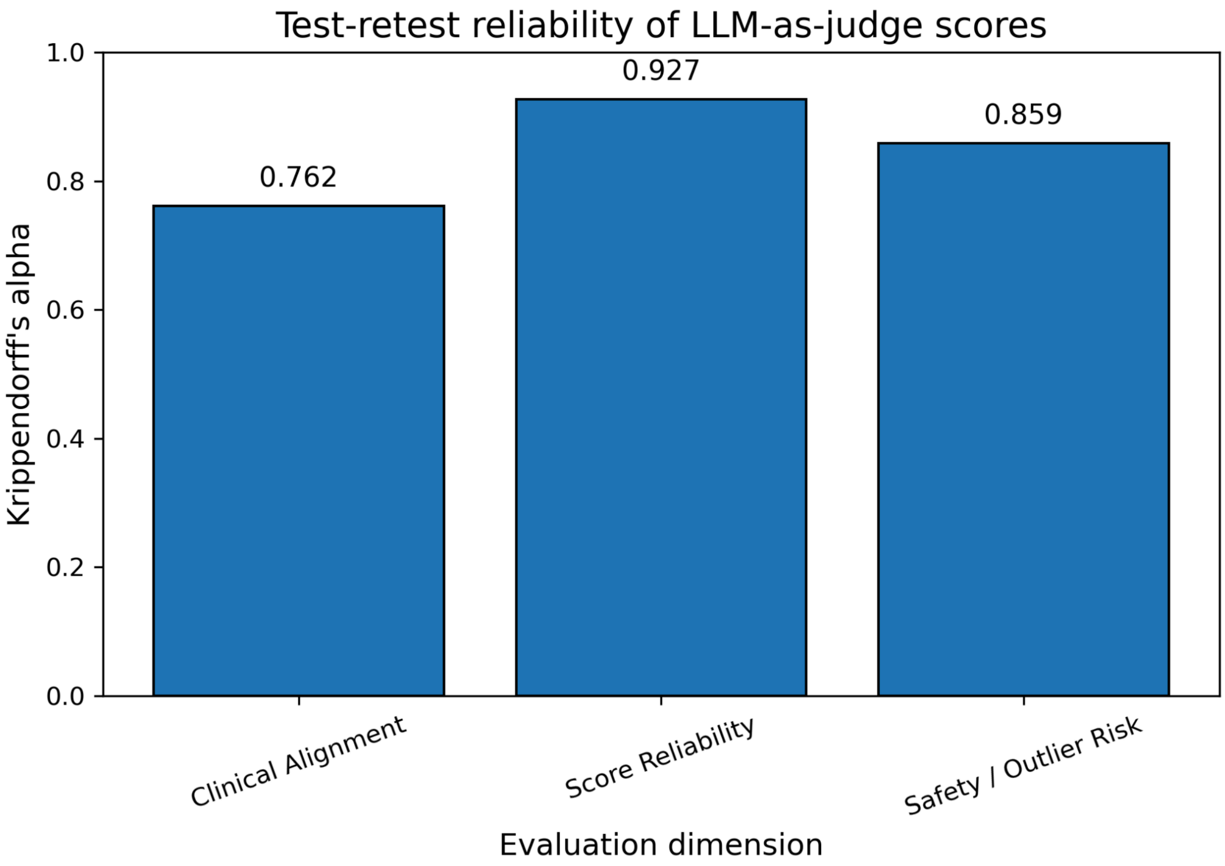


**Supplementary Figure 4: Test-retest reliability of the LLM-as-judge evaluation using Krippendorff's alpha.** Krippendorff's alpha was calculated across three repeated

LLM-as-judge runs for each fixed batch-model unit. Each unit was defined as one fixed patient batch evaluated for one scoring model. Reliability was assessed separately for clinical alignment, score reliability, and safety/outlier risk using interval-distance disagreement for the 1–5 Likert scores.
